# Understanding social and environmental factors and their contribution mechanisms to HIV transmission among heterosexual men in Indonesia

**DOI:** 10.1101/2022.03.15.22272326

**Authors:** Nelsensius Klau Fauk, Lillian Mwanri, Karen Hawke, Paul Russell Ward

## Abstract

The number of HIV infection among heterosexual men in Indonesia continues to increase. This paper describes social and environmental factors and the mechanisms through which these factors may have contributed to the transmission of HIV among men in Indonesia. A qualitative design using one-on-one and face-to-face in-depth interviews was employed to collect data from men living with HIV in Yogyakarta and Belu, from June to December 2019. Participants (n=40) were recruited using the snowball sampling technique. The logical model for socio-environmental determinants diagnosis was used to conceptualise the study and discuss the findings. The findings showed that social factors such as peer influence on sex, condom use and injecting drug use were contributing factors for HIV transmission among the participants. Other factors and drivers of HIV transmission included mobility, migration, and the environment the participants lived, worked and interacted, which facilitated their engagement in high-risk behaviours. The findings indicate the need for wide dissemination of information and education about HIV and condoms for men, within communities and migration areas in Indonesia and other similar settings globally in order to increase their understanding of the means of HIV transmission, and condom use for HIV prevention.

## Introduction

The 2021 UNAIDS report estimates 79.3 million people globally have been infected with HIV since the beginning of the epidemic, of which 36.6 million have died.^1^ The same report also estimates 37.7 million people were living with HIV (PLHIV) worldwide in 2020.^1^ Although new HIV infections continue to be reported every year, the global trend shows a decline in the number of annual new HIV infections from a peak of 2.9 million in 1997 to 1.5 million in 2020, which is a decline of 31% since 2010.^1,2^ In contrast to the global trend, the number of HIV cases in Indonesia has increased significantly during the same period, from 55,848 cases in 2010 to 191,073 cases in 2015 and 427,201 cases in 2021, and males represent 62% of total cases in the country.^3,4^

Previous studies exploring HIV transmission from the perspective of men living with HIV (MLHIV) have reported a range of HIV-risk factors. For example, high-risk behaviours, such as unprotected sex with multiple casual female partners or female sex workers (FSWs) and injecting drug use (IDU) practices, have been reported as proximal or direct factors that facilitate HIV transmission among men.^5-10^ Poor general knowledge about HIV and condoms for prevention is another contributing factor for HIV transmission among men as it leads to men being unaware of the possibility of contracting HIV through such high-risk behaviours they engage in.^11-13^ Men’s negative attitudes towards condom use, past negative experiences of condom use practices in previous sexual encounters,^12,14,15^ substance abuse and alcohol consumption prior to sex,^15-17^ and intention to avoid sex partner’s suspicion about their sexual relations with other women,^16,18^ are all supporting reasons for men’s engagement in unprotected sex. Peer influence, norms and pressures are also social factors that support men’s engagement in IDU, sex with multiple casual partners, and unprotected sex, which facilitate HIV transmission.^12,19-23^ The environment or communities in which men live and work, where brothels and illicit drugs are available and easily accessible, have also been reported as supporting factors for men’s engagement in unprotected sex with multiple casual partners,^5,21-23^ and IDU practices.^8,24,25^ A few studies have reported that relatively good financial condition or having money which enables the purchase of sexual services from women is also a contributing factor for HIV transmission among men as these studies have suggested that men who purchase sexual services from women are less likely to use condoms.^12^,^17^,^26-30^

Despite a range of HIV-risk factors among men reported globally, limited evidence exists in the context of heterosexual men in Indonesia. Previous HIV studies in the country have mainly focused on men who have sex with men (MSM),^19,22,31,32^ or have a limited focus on heterosexual men.^33,34^ Heterosexual contact and IDU practices among both men and women have been reported as the main behavioural risk factors for HIV transmission across the country, representing 70.1% and 7.9% of HIV transmission in the country respectively.^4^ These are evident by the prevalence among the young sexually active age groups; 70.7% of all infections among people aged 25 to 49, and 15.7% among the group aged 20 to 24.^4^ However, evidence about the mechanisms through which men are driven to engage in such high-risk behaviours for HIV transmission is still limited. This paper fills in these gaps in knowledge and adds further evidence on the social and environmental risk-factors, and the mechanisms through which these factors contribute to HIV transmission among men. It also extends our understanding of mobility and migration as drivers of HIV infection or as factors that drive men into risky social and environmental conditions which support their engagement in unprotected sex with multiple casual partners and IDU practices. Understanding these factors could be useful information for HIV programs or interventions that attempt to address HIV-risk factors and reduce HIV transmission among men in the study settings and any other similar settings in Indonesia and beyond.

## Methods

### Theoretical framework

The logical model for socio-environmental determinants diagnosis was used to conceptualise and discuss the findings of the current study.^35^ This model suggests that an individual’s behaviours are determined by other factors, such as knowledge, attitudes, beliefs, perceptions, and education. In the case of HIV infection, these factors may include a lack of knowledge of HIV and condoms, an inability to access condoms, negative personal attitudes and perceptions regarding condom use in relationships.^35-37^ Social networks, peer norms and influence, which lead to engagement in sex with multiple sex partners, unprotected sex and IDU practices are social factors that also determine an individual’s behaviours.^22,25,35,38,39^ The model also suggests that environmental factors are key determinants of health, and therefore multifaceted and dynamic nature of people’s environment should be considered in effort to understand their health behaviours and problems.^35,40^ Thus, it suggests that a health problem is determined by environmental factors that influence an individual’s behaviours or the behaviours of at-risk populations^35,41^ In the case of HIV problem, environmental factors or conditions that support people’s risky behaviours may include the conditions where sex work practices exist and illicit drugs are available and easily accessible.^35,37^

### Recruitment of the participants and data collection

This paper presents data from a large-scale qualitative study exploring the views or perspectives and experiences of PLHIV about HIV risk factors and impacts and their access to HIV healthcare services in Yogyakarta and Belu, Indonesia. The use of qualitative design was considered appropriate and effective when exploring participants’ perspectives and deep insight of their real-life experiences.^22,42^ It enabled the researchers to explore the participants’ stories, understandings and interpretations about the supporting factors for HIV transmission among them.

The recruitment of the study participants started with the field researcher (NKF) searching for assistance of the receptionists at HIV clinics in Yogyakarta and Belu to distribute the study information packs containing the field researcher’s contact details to potential participants or HIV patients who used their services. This was followed by the application of snowball sampling technique. PLHIV who called and stated their interest to participate in this study were recruited and scheduled for an interview based on their preferred time and place. Initial participants interviewed were also asked to distribute the information packs to their eligible friends and colleagues who might be willing to be interviewed about the topic being studies. This process was recursive and finally 40 men living with HIV (20 in Yogyakarta and 20 in Belu) aged 18 years old or above interviewed in the study. Only one participant withdrew his participation after the start of the interview for a few minutes due to personal reasons.

The participant’s age ranged from 22 to 60 years old, with the majority between 30 and 49 years. The majority of the men were married (n=25) and the others were unmarried. Nearly half of them (n=19) had been diagnosed with HIV for longer time, between 6 and 15 years. Several graduated from university, while the rest graduated from high school and elementary school. Sixteen participants reported having previously migrated and worked in other places in Indonesia or overseas.

Data collection was carried from June to December 2019 by way of one-on-one and face-to-face in-depth interviews. Interviews were conducted in a private room at the HIV clinic in Belu and in a rented house close to the HIV clinic in Yogyakarta where the initial information about the study was distributed. Interviews with the participants were audio recorded using a digital recorder, and fieldnotes were also undertaken by the researcher during each interview once felt necessary. Interview duration varied between 35 to 87 minutes. Interviews were focused on several main areas, including participants’ perceptions about factors that have facilitated HIV transmission among them. Interviews also explored social factors, such as peer influence, norm, pressure, and how these factors contributed to their risky behaviours or influence their sexual relations, practices, condom use behaviours, engagement in IDU, and HIV transmission among them. Environmental conditions or communities where the participants moved into, lived, worked, interacted, and how these factors influence their engagement in IDU, sexual relations, sexual practices, condom use behaviours, and contributed to HIV transmission among them, were also explored. The interviews also explored factors that drove the participants into risky social and environmental conditions that facilitated their engagement in such risky behaviours for HIV transmission. Recruitment and data collection ceased when the researchers felt that information provided by the participants had been rich enough to answer the research questions and objectives or data saturation had been reached. Data saturation was decided when the last few participants in each setting seemed to provide information similar to that of previous participants. Interviews were conducted in Bahasa, the primary language of the field researcher and the participants.

### Data analysis

Data analysis started with the transcription of the audio recordings of the interviews performed manually. The transcripts were then imported to NVivo 12 where the comprehensive analysis was performed. The five steps of qualitative data analysis introduced in Ritchie and Spencer’s analysis framework was used to guide this data analysis process.^43^ These included: (i) familiarisation with the data, (ii) identification of a thematic framework, (iii) indexing the data, (iv) charting the data and (v) mapping and interpreting the data. These steps helped the management of these qualitative data in a coherent and structured way, and enhanced the analytic process in a rigorous, transparent and valid way. The entire data analysis was carried out in Bahasa to retain the social meanings of the information from the participants.^44^

Ethics approval was obtained from the Social and Behavioural Research Ethics Committee, Flinders University (No. 8286).

## Results

### Social factors and HIV transmission

#### Peer influence on sex and condom use practices

Across both settings, peer influence was reported as a supporting factor for the engagement of participants in sex with multiple partners, including with FSWs, a practice that facilitated HIV transmission. A mechanism of peer influence was reflected in the acts of inviting each other to hang out and look for FSWs during weekends or after working hours. Such an influence seemed to be supported by the cohabitation conditions of work areas or construction sites which facilitated close relationships between peers or fellow workers. These are illustrated in the following narratives of two married participants who lived in plantation areas or construction sites within or outside of Indonesia, together with other workers from different countries for several years

> *“We (the man and his male friends) were from different countries and worked together (he worked abroad in oil palm plantation). During the weekend my friends often asked me to hang out to look for the girls (either female migrant workers or FSWs) to have sex with. Finally, I felt accustomed to it and every weekend we often went out to have sex” (MP10, Yogyakarta)*.

> *“Construction work sometimes takes months and years to finish and we (construction workers) are all men. We do not work at the weekend, and sometimes we invite each other to hang out and look for girls (FSWs). Influencing each other for this (sex with FSWs) always happens in the construction area because we live together” (MP3, Belu)*.

The same story was also shared by non-married participants from both study settings as reflected in the following narrative of a non-married man who acknowledged living together with peers who had already been involved in sex with FSWs as an influencing factor for his prolonged engagement in such practice:

> *“At that time (he worked in another province in Indonesia, for several years) I lived together with my friends who liked to ‘play women’ (have sex with FSWs). So, if one of us wanted, then he would invite others to go together. I engaged in such practice for five years of working period over there” (MP5, Belu)*.

Peer influence was also a factor that supported participants’ engagement in unprotected sexual behaviours, which facilitated HIV transmission among them. The influence was reflected in various discouraging words for condom use as shown in the following narrative of participants who had heard of condoms prior to their HIV diagnosis:

> *“I have heard about condoms when I was working in XX (name of another province in Indonesia) but I never used them every time I visited those girls (had sex with FSWs) because my friends said that it hurts and makes you feel uncomfortable during the sex” (MP20, Belu)*.

> *“I know about condoms (before he was diagnosed with HIV), but I have not started using condoms because my friends said using condoms make it (sexual intercourse) less pleasurable. I just believed in what they said and never tried to use a condom every time I had sex” (MP2, Yogyakarta)*.

#### Peer influence on injecting drug use practices

Peer influence was indicated as playing a supporting role in the engagement of the participants in IDU. Yogyakarta participants who reported infection through IDU practices described they were initially introduced to injecting drugs by their friends, and engaged in such practices together with them, a factor which was not identified among participants in Belu. Introduction of injecting drugs by peers and peer invitation to use drugs together were some mechanisms through which social factors (e.g., peer influence) influenced the participants’ behaviours and contributed to HIV transmission among them:

> *“I used to use injecting drugs together with my friends, we were close friends. We used drugs every time we got together at our meeting spots. …. We always invited each other to use (drugs together). …. I got to know about illicit drugs through social relations, through my friends. They were the ones who introduced injecting drugs to me at the first time” (MP1, Yogyakarta)*.

Purchasing drugs together with friends, who were also drug users, was another mechanism through which social factors (e.g., peer influence) supported the participants’ continuous engagement in illicit drug use that had facilitated HIV transmission among them. The participants described that they and their friends, or other drug users, often had to contribute some amount of money to buy drugs together when they could not afford to buy drugs individually. This seemed to be a common strategy used by the participants to support their engagement in IDU practices as illustrated in the following narratives of participants who engaged in IDU for many years prior to their marriage:

> *“…. My friends and I contributed (some amount of money) together. At that time (when he was in senior high school), twenty thousand rupiahs could be used to buy (drugs) which could be used two or three times” (MP13, Yogyakarta)*.

> *“In the prison, if we did not have enough money then we bought the drugs together and then used together” (MP12, Yogyakarta)*.

The engagement of these participants in IDU seemed to also be influenced by the social situations in the places where they lived. Some participants described that illicit drugs were popular at the time they started getting involved in drug use practices, and using drugs made them ‘feel’ popular among their friends. These were reflected in the following quote from a participant who acknowledged acquiring HIV through IDU and living with HIV for more than ten years:

> *“I used heroin for three years, injecting drug. At the time I was (studying) at the university in Bandung, heroin was so popular. …. Back in those time, getting drunk and using drugs made you feel cool and popular among friends. I used to get together with friends from wealthy families and at the beginning they often bought the drugs and we used together” (MP11, Yogyakarta)*.

### Environmental factors and HIV transmission

#### The influence of environmental factors on sexual behaviours

The environment surrounding participants was also reported as playing an important role in supporting their engagement in sex with multiple sex partners, or changing sex partners over time. For example, the availability of brothels and FSWs within workplace environments in which men and women lived together, such as plantation areas, were described by the participants across both study settings:

> *“There were many girls (FSWs) from around the world over there (Thailand. He worked in Thailand for several years). It (Thailand) is like the centre, they (FSWs) were from around the world. So, it was easy for me to find them and I just needed to choose the ones I liked” (MP4, Belu)*.

> *“I often had sex with different women at the workplace (he used to work at oil palm plantation in Malaysia) because there were many female workers, and many were widowed. …. I often had sex with different women because the environment was supportive. I could meet women (female workers) from different countries, and we, both women and men, stayed together in the plantation area” (MP10, Yogyakarta)*.

Some men also made a similar acknowledgement of the availability of brothels and FSWs within communities where they lived as an environmental factor for their engagement in sex with multiple FSWs. The following story from a participant who had been diagnosed with HIV for many years reflects such influence on his sexual behaviour since he was in high school:

> *“There was a brothel in the place where I lived I had sex with some of the girls (FSWs). …. I realised that environmental factor had a big influence on my engagement in sex with them. When I was still in senior high school, my house was close to the brothel, only one kilometer, and every night the girls stood on the side of the road. Clients could choose any girls they liked. That was the time I initially engaged in sex and after that I often had sex with them (FSWs)” (MP9, Yogyakarta)*.

#### The influence of environmental factors on injecting drug use practices

Environmental factors were also reported to have an influence on IDU practices that facilitated HIV transmission among the majority of participants in Yogyakarta, which were not identified in the interviews with participants in Belu. Some participants reported illicit drugs were easily available and accessible where they lived, and was one of the supporting factors for their engagement in IDU practices through which they acquired HIV:

> *“I used injecting drugs for nine years. The place where I used to live was called drug village. People sold drugs in every alley. So, there were people who distributed drugs every day. It was not difficult to find drugs” (MP14, Yogyakarta)*.

The workplace environment which was far away from family was another supporting factor for the use of injecting drugs among the participants. This was due to the absence of supervision and restrictions from parents and other family members at the places where the participants worked and lived. Such absence was described as supporting their involvement in the use of illicit or injecting drugs, as they were not afraid of anybody and were not being watched by family members or other people around them:

> *“It was free (to use drugs) at the place where I was busking, there were not my parents or any other family members. So, I was not afraid of anybody. There were only my friends who were also (drug) users. After using (injecting drugs) I was busking and after busking if we (the man and his friends) still had (drugs) then we again injected” (MP3, Yogyakarta)*.

Similarly, the place where participants were detained was also an environmental factor that supported their engagement in IDU practices. A few participants described that the prison environment was very supportive of drug use due to the availability and accessibility of drugs. This was indicated as supporting the transmission of HIV infection among them due to the practice of needle sharing and non-disclosure of their HIV status among drug users. The following narrative of a participant who admitted to have been detained in prison twice due to drug abuse reflects this claim:

> *“Never run out of stock (of drugs) in the prison. You can use whatever (drugs) you want in the prison, they are available. The environment was very supportive, drugs were available and easily accessible, thus I used it nearly every day. …. Every night, there was music in every room just like in a discotheque. It was highly likely to get HIV in prison if we did not have information about it. Some deliberately spread it (HIV) because we shared the same needles. …. We did not know who was infected. It was not possible to ask about it, while the ones who were (HIV) positive did not want to disclose their status due to the fear discrimination in prison” (MP13, Yogyakarta)*.

The surroundings or conditions where clean needles were not readily available and difficult to access were also environmental factors that supported the needle-sharing practice among injecting drug users, which facilitated HIV transmission among them. All the participants who acknowledged contracting HIV through IDU, described how needles or injecting devices were limited and difficult to buy back in the 90s and early 2000s, which led to high-risk injecting drug behaviour and HIV transmission:

> *“I shared needle (with other drug users) because it was difficult to buy a new one. No pharmacies could give you needles if you wanted to buy. So, sometimes I sharpened the syringes that were no longer sharp, and we reused them together” (MP11, Yogyakarta)*.

#### Mobility and migration as drivers of HIV transmission

Mobility within Indonesia for work purposes was indicated to play an important role in the transmission of HIV among the participants. It was indicated to drive the participants into risky social and environmental conditions that influenced their behaviours or supported their engagement in unprotected sex and IDU practices that facilitated HIV transmission. This is illustrated in the following quote from a participant:

> *“I was a construction worker for so many years, so I moved from one place to another, worked and lived with my colleagues for months. Most of my them looked for girls (FSWs) every weekend, that was the situation which led me to engaging in sex with sex workers and got infected with HIV” (MP3, Belu)*.

Similarly, participants across both study settings reported migration for work purposes within or outside of Indonesia was also a driver of HIV transmission, supporting engagement in unprotected sex with multiple casual female partners and IDU practices, and facilitating HIV transmission among them:

> *“I used injecting drugs for the first time in XX (name of a place where he worked). At that time, I moved over there and worked at oil palm plantation for a year. I had many friends at the plantation who were injecting drug users and I used injecting drugs too. We were alone, nobody cared about us, and we were not afraid of anybody. Many of us used drugs, so it was obvious that we influenced each other. So, drugs and women (having sex with casual female partners or FSWs) were two things that I could not avoid. Many of us (men) did these. I was away from my wife and families, so I just did those things” (MP5, Yogyakarta)*.

## Discussion

This paper presents an understanding of risk factors for HIV transmission among heterosexual men in Yogyakarta and Belu, Indonesia. The findings suggest that peer influence impacted participants’ behaviours through various mechanisms (e.g., invitation and encouragement for sex with multiple casual female partners, discouragement towards condom use, etc) which played an important role in the transmission of HIV among them. These peer influences were compounded by mobility and migration for work purposes, in which the participants found themselves in situations where high risk behaviour was normalised. Physical distance from partners, spouses or girlfriends for long periods of time, meeting and socialising with new people from different places, easy access to FSWs, and limited education about condom use, facilitated an environment of casual and unprotected sex. These findings add to literature about the concept of social influence in the logical model^35^ and to findings about peer influence as reported in previous studies involving different groups of men, such as clients of FSWs, motorbike taxi drivers and men who have sex with men in Indonesia.^21-23^ These previous studies have reported peer influence through connecting each other to and sharing information about casual sex partners or FSWs, as a social factor that increased the frequency of their engagement in sex with multiple casual partners and inconsistent condom use practices.

Peer influence through introduction of illicit/injecting drugs, including purchasing and using injecting drugs together, also facilitated HIV transmission, adding to current knowledge about factors associated with IDU practices, such as socio-structural factors (e.g., social networks or interactions, socialisation into drug-using identity, etc) and individual factors (e.g., seeking pleasure, coping with pain or trauma, etc).^8,9,25,45^ Social perception of drug use as a factor that increased one’s popularity, was also a factor that motivated some participants’ involvement in IDU practice, which has not been explored in previous studies on this topic.^8,25,45^ Peer influence on IDU was not a factor identified in the interviews with participants in Belu, which seemed to be reflective of their unawareness of illicit drugs and an indication of the unavailability of illicit drugs in their social relationships.

The environment in which the participants lived and worked, where brothels and FSWs were available, and which allowed men and women to live together, such as in plantation areas, was also a supporting factor for their engagement in sex with multiple casual female partners and FSWs. For some men across the study settings, their mobility and migration for work drove them into such environments, which supported their engagement in risky sexual behaviour. These current findings add further evidence to the findings of previous studies,^5,21-23,46,47^ which have reported environmental conditions that support access to brothels or FSWs, an establishment of sexual networks, and social influence as facilitators of men’s engagement in unprotected sex with multiple casual partners, which puts them at high risk for HIV acquisition.

The current findings also suggest that the surroundings (e.g., community where the participants lived, workplaces, and prison) where illicit drugs were available and accessible, and clean needles were not readily available and difficult to access, led to being infected with HIV. The findings extend our understanding about environmental-related risk factors associated with the practice of IDU, such as family structure, parental drug use, homelessness or unstable housing, and availability of drugs within communities, as have been reported the previous studies.^9,25,45,48,49^ The stories of participants in Yogyakarta illustrated how some risky environments where they moved into, lived and worked had played a pivotal role in their HIV acquisition. For example, working sites in migration areas and busking spots where there were a lack of supervision and restriction from families and significant others, were environmental factors that encouraged their self-confidence and engagement in IDU practices. The findings conform to the constructs of the logical model^35^ which suggest that environmental factors (e.g., in this study: brothels, working sites in migration areas and busking spots) influence people’s behaviours (e.g., in this case: facilitate participants’ engagement in IDU), which lead to health problems (e.g., HIV infection). The findings also reflect the influence of the environment or circumstances where people live, work and grow on their health and well-being, which has been a core element in the concept of social determinants of health.^50^

## Conclusions

Social and environmental factors play a pivotal role in HIV transmission among heterosexual men in Indonesia. These include peer influence on participants’ sexual behaviours and IDU practices, and communities or areas that participants live and/or work in, where brothels, FSWs, and illicit drugs are often available and easily accessible. Mobility and migration to other places for work purposes are part of these drivers leading to HIV transmission. The findings indicate the need for better education about the prevalence of HIV and how it is transmitted, and accurate information about condoms, especially for heterosexual males in high-risk settings (working away from home for example), Future large-scale studies to understand men’s perceptions on various HIV-risk factors are recommended.

## Data Availability

All data produced in the present work are contained in the manuscript.

## Acknowledgements

We would like to thank the participants who had spent their time to voluntarily take part in the interview and provided us with valuable information.

